# Mendelian Randomization Study of Whole Blood Viscosity and Cardiovascular Diseases

**DOI:** 10.1101/2023.10.30.23297662

**Authors:** Youngjune Bhak, Albert Tenesa

## Abstract

**Aims:** Association between whole blood viscosity (WBV) and an increased risk of cardiovascular disease (CVD) has been reported. However, the causal relationship between WBV and CVD remains not thoroughly investigated. The aim of this study was to investigate the causal relation between WBV and CVD.

**Methods:** Mendelian randomization was employed to investigate the casual relationship between WBV and CVD. Calculated WBV, derived polygenic risk score (PRS) for WBV, and medical records from 378,210 individuals participating in the UK biobank study were analyzed.

**Results:** The means of calculated WBVs were 16.9 (standard deviation: 0.8) and 55.1 (standard deviation: 17.2) for high shear rate (HSR) and low shear rate (LSR), respectively. 37,859 (10.0%) major cardiovascular event (MACE) consisted of 23,894 (6.3%) cases of myocardial infarction (MI), 9,245 (2.4%) cases of ischemic stroke, 10,377 (2.7%) cases of revascularization, and 5,703 (1.5%) cases of Coronary heart disease-related death. WBVs of individuals with PRS above median were significantly higher than the group that were below median for both low shear rate (P<0.001) and high shear rate (P<0.001). The odd ratios (95% confidence intervals) for MACE in the higher WBV group were 1.22 (1.19 - 1.25) and 1.14 (1.12 - 1.16), for HSR and LSR, respectively. The higher PRS group was insignificant with all outcomes, for both HSR and LSR.

**Conclusions:** The Mendelian randomization analysis conducted in this study does not support a causal relationship between calculated WBV and CVD.

## Introduction

Cardiovascular diseases (CVD), including heart attack and stroke, are one of leading causes of morbidity and mortality globally [1]. Association between the risk of CVD and Whole blood viscosity (WBV), a measure of the thickness and flow resistance of bulk blood, have been reported [2-5]. However, establishing a causal relationship between WBV and CVD remains challenging due to the potential biases from confounding factors in traditional studies lacking randomized trial designs.

Mendelian randomization (MR), an epidemiological method, utilizes genetic variants robustly associated with an exposure of interest as instrumental variables (IVs) to investigate the causal effects of risk factors on specific outcomes [6]. The advantage of MR lies in the random assignment of these genetic variants at conception, rendering MR studies less susceptible to confounding factors compared to traditional observational studies [7]. Furthermore, MR is robust against reverse causality since the development of diseases does not alter individuals’ genotypes. MR have been utilized to investigate casual relationships of risk factors such as blood pressure [8, 9], obesity [10], type 2 diabetes mellitus [11] and, profile of blood lipids [12-14] in and CVD.

The objective of this study was to use MR to examine the causal relationship between WBV and CVD. WBV of individuals was calculated by applying the formula previously reported [15]. Polygenic risk score (PRS), a quantitative genetic risk score, representing the cumulative impact of genetic variants, was derived and used as the instrumental variable in MR. Causal relationship of WBV on risk of CVDs were assessed by comparing individuals stratified by their derived PRS.

## Methods

### Participants

The UK Biobank (UKB) is a prospective research resource of population-based cohort study that include comprehensive phenotype and genotype data from approximately 500,000 participants recruited in 2006 - 2010 residing in England, Scotland, and Wales (www.ukbiobank.ac.uk). This an open-access resource was established to support investigations into the factors influencing various health outcomes [16].

### Ethics Statement

The UK Biobank project was approved by the National Research Ethics Service Committee North West-Haydock (REC reference: 11/NW/0382). Participants provided written informed consent to participate in the UK Biobank. An electronic signed consent was obtained from the participants. This research was conducted using the UK Biobank Resource under project 44986.

### Extrapolation of Whole Blood Viscosity

WBV was calculated for both low shear rate (LSR) (0.5 sec^-1^) and high shear rate (HSR) (208 sec^-1^) from hematocrit (HCT) and total plasma protein concentration (TP) using the validated formulation [15]. HCT was calculated by multiplying red blood cell count by the mean corpuscular volume.

HSR: WBV (208 sec^-1^) = (0.12 × HCT) + (0.17 TP) - 2.07

LSR: WBV (0.5 sec^-1^) = (1.89 × HCT) + (3.76 TP) - 78.42

### Study outcomes

The data pertaining to each component of the participants’ outcomes in the present study was accessible through the UK Biobank study [16]. The primary outcome of the study was major cardiovascular events (MACE), which encompassed a composite outcome involving the occurrence of non-fatal MI, coronary revascularization (defined as “percutaneous transluminal coronary angioplasty, PTCA” or “coronary artery bypass grafting, CABG”), ischemic stroke, or death due to coronary heart disease (CHD).

These specific outcomes were defined and categorized as follow: non-fatal MI defined algorithmically by UK Biobank (ICD9: 410.X, 411.0.X, 412.X, 429.79; ICD10: I21.X, I22.X, I23.X, I24.1, I25.2; self-report 20002: 1075), PTCA or CABG (self-report 20004: 1070, 1095, 1523; Procedures (OPCS): K50.1, K40.X, K41.X, K42.X, K43.X, K44.X), ischemic stroke (ICD9: 434.X, 436.X; ICD10: I63.X, I64.X; self-report 20002: 1583), and death due to CHD (Death 40001, 40002: I21.X, I22.X, I23.X, I24.X, I25.1, I25.2, I25.3, I25.5, I25.6, I25.8, I25.9) [14].

### Genetic instrument

We conducted a split-sample genome wide association study (GWAS) and MR using the UK Biobank dataset. The dataset was randomly split into two halves, and for each half, GWASs for WBVs was performed. The resulting summary statistics from each half were then used to derive the PRS of WBV (WBV-PRS) for the other half to avoid sample overlap [17]. To ensure homogeneity, we limited our analyses to unrelated individuals of White British ancestry. Additionally, individuals with more than 10% missing genotypes or those with discrepancies between recorded sex and genetically determined sex were excluded. Following these exclusions, the final dataset consisted of 378,213 participants (Table 1).

GWASs were conducted using genotypes of the unrelated White British individuals. White British individuals are inferred from UK Biobank records. The unrelated individuals were identified using the KING software with following options: --unrelated --degree 2 (version 2.2.8) [18]. Autosomal genotypes of unrelated White British were further filtered using PLINK software (version 1.90p) with the following options; --geno 0.01, --hwe 1e-15, --maf 0.01, and --mind 0.1 [19]. For the GWASs of WBVs, REGENIE software (version 3.2.2) was utilized with the following option; --apply-rint [20]. Covariates considered in the GWAS included age, age square, genetic principal components 1 to 20, sex, and genotyping array.

**Table 1.** Baseline characteristics.

| Table 1. Baseline Characteristics of the Participants.* |  |
| --- | --- |
| Variable | Participants<br>(N=378,210) |
| Age (yr) | 57.0 ± 7.9 |
| Female sex (%) | 53.7 |
| Blood pressure (mm Hg) |  |
| Systolic | 140.3 ± 19.7 |
| Diastolic | 82.3 ± 10.7 |
| Body-mass index | 27.4 ± 4.7 |
| Diabetes (%) | 4.8 |
| Current smoker (%) | 10.1 |
| Lipid levels (mg/dl) |  |
| Total cholesterol | 221.0 ± 44.3 |
| LDL cholesterol | 138.1 ± 33.7 |
| HDL cholesterol | 56.2 ± 14.8 |
| Triglyceride level | 155.8 ± 90.6 |
| Whole blood viscosity |  |
| LSR | 55.1 ± 17.2 |
| HSR | 16.9 ± 0.8 |
| Outcomes (%) |  |
| Major cardiovascular event | 37,857 (10.0) |
| Myocardial infarction | 23,893 (6.3) |
| Ischemic stroke | 9,245 (2.4) |
| Revascularization | 10,376 (2.7) |
| Death due to CHD | 5,703 (1.5) |

- 1 Plus-minus values are means ±SD. HDL denotes high-density lipoprotein, LDL low-density lipoprotein, LSR - 2 low-shear rate, HSR high shear rate, and CHD coronary heart disease. Major cardiovascular event includes - 3 myocardial infarction, ischemic stroke, revascularization, and death due to CHD

To select independent instrumental variants for Mendelian randomization, summary statistics were clumped to extract index variants using PLINK software with the following options; -- clump, --clump-p1 0.00000005, --clump-r2 0.001, and --clump-kb 10000 [19]. To avoid the risk of weak instrument bias, variants with F-statistics > 10 were selected [21]. The variants were filtered out if a variant had a reported association with CVD and or factors related with blood viscosity or CVD. The associations were investigated by utilizing PhenoScanner database with the following options; catalogue: diseases & traits, p-value: 5×10^−8^, proxies -EUR, r^2^: 0.8, and build: 37 [22, 23]. Such factors include obesity [10, 24-26], blood pressure [8, 9, 27], lipid traits [12-14, 28-32], type 2 diabetes mellitus [11, 33, 34], smoking and alcohol intake [35-37]. For HSR, 53 variant from one half split and 50 variants from the other half split passed the filters, respectively. For LSR, 58 variant from one half split and 52 variants from the other half split passed the filters, respectively (Table S1-S4). The filter passed variants were used to derive WBV-PRS. The WBV-PRS was derived using PRSice-2 software (version 2.3.3) by using the summary statistics of variants that passed our filters [38].

## Statistical analyses

Categorical variables were presented as counts and percentages, continuous variables were presented as mean and standard deviations (SD). Odd ratio (OR) with 95% confidence interval (CI) were analysed using logistic regression. The analysis with WBV-PRS was accompanied by adjustments for the split halves, considering them as a batch effect. All significance tests were two-tailed, and statistical significance was determined at p < 0.05. The statistical analyses were performed using the R version 4.2.1 (R Foundation for Statistical Computing, Vienna, Austria).

## Results

A total of 378,210 individuals were included in the study (Table 1). The mean age ± SD was 57.0 ± 7.9 years. 53.7% of participants were women. The mean ± SD of calculated WBV were 55.1 ± 17.2 and 16.9 ± 0.8 for LSR and HSR, respectively. 37,859 (10.0%) of major cardiovascular events, including 23,893 (6.3%) myocardial infarction, 9,245 (2.4%) ischemic stroke, 10,376 (2.7%) revascularization, and 5,703 (1.5%) death due to CHD were recorded. A group of individuals with a WBV-PRS above the median (High WBV-PRS) have higher WBV compared to those with a WBV-PRS equal to or below the median for both HSR (mean difference: 0.11, P<0.001) and LSR (2.58, P<0.001).

Associations of the WBV and WBV-PRS groups with the risk of CVDs have exhibited differences. Compared to a group of individual with HSR equal to or below the median, a group of individuals with HSR above the median showed significant association with the increased risk of CVDs including MACE (Odd ratio: 1.22, 95% Confidence interval: 1.19 - 1.25, P<0.001), MI (1.23, 1.20 - 1.27, P<0.001), stroke (1.08, 1.03 - 1.12, P<0.001), revascularization (1.29, 1.23 - 1.34, P<0.001), and death by CVD (1.30, 1.122 - 1.37, P<0.001). High HSR-PRS was not significant in all CVD categories for both unadjusted and adjusted study (Fig. 1A).

**Figure 1.**
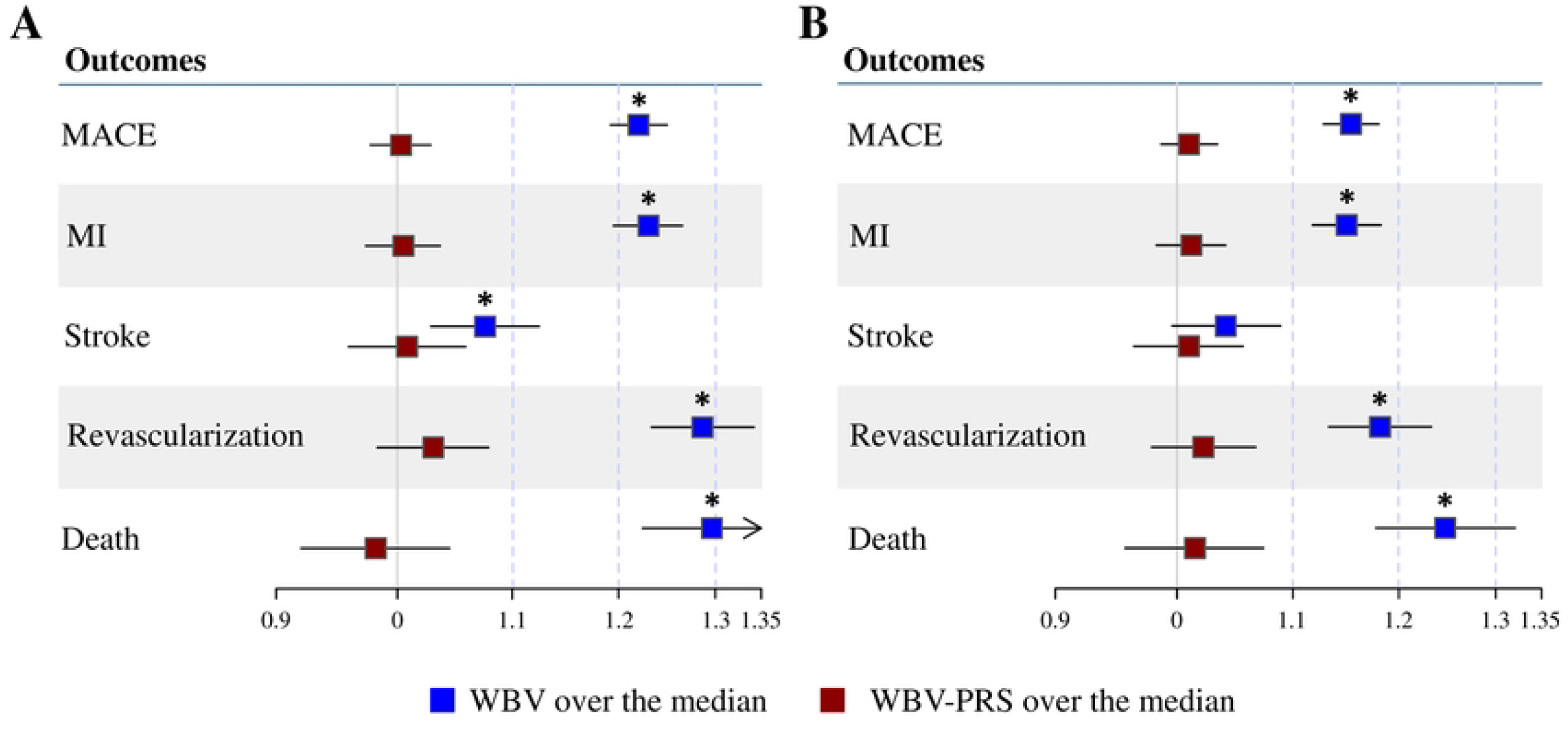
Association of the WBV and WBV-PRS with the risk of cardiovascular events. Panel A shows the associations between high HSR and high HSR-PRS with the risk of cardiovascular events, and panel B shows the associations between high LSR and high LSR-PRS with the risk of cardiovascular events. The boxes represent point odd ratio estimates of the effect, and the lines represent 95% confidence intervals. MACE denotes major cardiovascular event, MI myocardial infarction, WBV whole blood viscosity, WBV-PRS polygenic risk score for whole blood viscosity.

Compared to a group with LSR equal to or below the median, The group with LSR above the median showed a significant association with the increased risk of CVDs including MACE (1.15, 1.13 - 1.18, P<0.001), MI (1.15, 1.12 - 1.18, P<0.001), revascularization (1.18, 1.13 - 1.23, P<0.001), and death by CVD (1.25, 1.18 – 1.32, P<0.001). Effect of High LSR-PRS was insignificant in all CVD for both unadjusted and adjusted study (Fig. 1B).

## Discussion

This study used MR to investigate the potential causal association between WBV and CVD. The result from this study indicated insufficient evidence to substantiate a causal link between WBV and the risk of CVD.

WBV is susceptible to various influencing factors. Notably, WBV demonstrate non-Newtonian fluid behaviour. Under conditions of low shear rates, blood cells tend to aggregate, leading to an elevation in viscosity. Conversely at HSR, the opposite phenomenon occurs [39-41]. WBV has been noted to exhibit association with CVD and cardiovascular risk factors in previous studies [3, 42, 43]. However, upon adjustments for these risk factors, the association between WBV and CVD was found to be statistically non-significant in the subsequent study [44]. The not significant association following adjustments remained consistently evident in this study using MR.

However, the study result should be interpreted and considered carefully since we have not utilized directly measured WBV. While the formula employed in our study has undergone validation and has been applied in previous researches [15, 45, 46], it does not considered factors for WBV such as blood cell aggregability and deformability [47]. To establish a robust causal link between WBV and both CVD and CVD-related factors, future studies should incorporate measured WBV values, taking into account these critical variables.

## Data Availability

Our data is available as a part of the UK Biobank project. Details of procedures for accessing the UK Biobank data can be found here: https://www.ukbiobank.ac.uk/enable-your-research/apply-for-access

## Acknowledgments

This work used the Edinburgh Compute and Data Facility (ECDF) (http://www.ecdf.ed.ac.uk/).

This research has been conducted using the UK Biobank Resource project 44986. For the purpose of open access, the author has applied a CC-BY public copyright licence to any Author Accepted Manuscript version arising from this submission.

## Authors’ contributions

Conceptualization: Y.B.

Data curation: Y.B.

Formal analysis: Y.B.

Funding Acquisition: A.T.

Investigation: Y.B.

Methodology: Y.B.

Project Administration: Y.B., A.T.

Resources: Y.B.

Software: Y.B.

Supervision: A.T.

Visualization: Y.B.

Writing – original draft: Y.B.

Writing – review & editing: Y.B., A.T.

